# Immunogenicity and safety of an inactivated SARS-CoV-2 vaccine in people living with HIV-1

**DOI:** 10.1101/2021.09.14.21263556

**Authors:** Yanmeng Feng, Yifan Zhang, Zhangyufan He, Haojie Huang, Xiangxiang Tian, Gang Wang, Daihong Chen, Yanqin Ren, Liqiu Jia, Wanhai Wang, Jing Wu, Lingyun Shao, Wenhong Zhang, Heng Tang, Yanmin Wan

## Abstract

**Background:** It has been proven that inactivated COVID-19 vaccines are safe and effective in general population with intact immunity. However, their safety and immunogenicity have not been demonstrated in people living with HIV (PLWH).

**Methods:** 42 HIV-1 infected individuals who were stable on cART and 28 healthy individuals were enrolled in this study. Two doses of an inactivated COVID-19 vaccine (BIBP-CorV) were given 4 weeks apart. The safety and reactogenicity of the vaccine were evaluated by observing clinical adverse events and solicited local and systemic reactions. Humoral responses were measured by anti-spike IgG ELISA and surrogate neutralization assays. Cell-mediated immune responses and vaccine induced T cell activation were measured by flow cytometry.

**Findings:** All the HIV-1 infected participants had a CD4^+^ T cell count of above 200 cells/μL both at baseline and 4 weeks after vaccination. No solicited adverse reaction was observed among all participants. Similar binding antibody, neutralizing antibody and S protein specific T cell responses were elicited in PLWH and healthy individuals. Further analyses showed that PLWH with low baseline CD4^+^/CD8^+^ T cell ratios (<0·6) generated lower antibody responses after vaccination than PLWH with medium (0·6∼1·0) or high (≥1·0) baseline CD4^+^/CD8^+^ T cell ratios (P<0·01). The CD3^+^, CD4^+^ and CD8^+^ T cell counts of PLWH decreased significantly after vaccination, but it did not lead to any adverse clinical manifestation. Moreover, we found that the general burden of HIV-1 among the PLWH cohort decreased significantly (P=0·0192) after vaccination. And the alteration of HIV-1 viral load was not significantly associated with the vaccine induced CD4^+^ T cell activation.

**Interpretation:** Our data demonstrate that the inactivated COVID-19 vaccine is safe and immunogenic in PLWH who are stable on cART with unsuppressed CD4 counts.

**Funding:** This work was funded by the National Natural Science Foundation of China (Grant No. 81971559, 82041010).

**Research in context:** *Evidence before this study:* The safety and efficacy of inactivated COVID-19 vaccines have been validated in general population with intact immunity. However, their safety and immunogenicity have not been demonstrated in people living with HIV (PLWH).

*Added value of this study:* Our study provides the first evidence to show humoral and cellular immune responses to an inactivated vaccine in PLWH who have been stable on cART with good CD4 cell counts. We found that participants with HIV-1 generated antibody and T cell responses comparable with those of healthy individuals after two-dose vaccination. The baseline CD4/CD8 ratios while not the absolute CD4^+^ T cell counts were shown to be associated with the magnitudes of vaccine induced antibody responses. Moreover, we showed that the vaccine induced T cell activation did not increase the viral burden in PLWH on cART. On the contrary, the levels of plasma HIV-1 RNA decreased among a significant percentage of PLWH.

*Implications of all the available evidence:* Our data demonstrate that the inactivated COVID-19 vaccine is safe and immunogenic in PLWH who are stable on cART with unsuppressed CD4 counts and indicate that this vaccine might be protective and efficacious against COVID-19 for people with HIV.

## Introduction

Even though the well-controlled HIV infection per se is not found as a risk factor of increased SARS-CoV-2 prevalence,^1,2^ it is alerted that the compromised immunities and the high frequencies of comorbidities may render excessive challenges to this population.^3,4^ A more recently released WHO report (WHO reference number: WHO/2019-nCoV/Clinical/HIV/2021.1) suggests that HIV infection appears to be a significant independent risk factor for both severe/critical COVID-19 presentation at hospital admission and in-hospital mortality. Vaccination against SARS-CoV-2 for PLWH has also been recommended by WHO and health authorities of many countries. However, as a clinical observation showed that SARS-CoV-2 natural infection induced lower protective antibody responses in people with HIV,^5^ it is concerned that the immunogenicity of COVID-19 in PLWH might be weak. The anxiety escalated when the news released from the Novavax COVID-19 vaccine study suggested that HIV infection might dampen the vaccine effectiveness.^6^

Reassuringly, more recently published data showed that both the messenger RNA vaccine and the ChAdOx1 nCoV-19 vaccine could elicit protective antibody responses in PLWH comparable with those in healthy individuals.^7-10^ In this study, we compared the antibody responses to two-dose inactivated SARS-CoV-2 vaccination between healthy vaccinees and HIV-1 infected individuals stable on cART. We demonstrated that the inactivated COVID-19 vaccine was immunogenic and safe in PLWH. Baseline CD4^+^/CD8^+^ T cell ratios were found to be associated with both the RBD binding antibody and the neutralizing antibody responses in PLWH.

## Materials and methods

### Study design and participants

In this open-label two-arm non-randomized study, we enrolled a cohort of HIV-1 infected individuals (n=42) who were stable on cART and under routine follow-up at Hubei Provincial Center for Disease Control and Prevention, China and a cohort of healthy individuals (n=28). Written informed consent was obtained from all participants, and the study was approved by the Research Ethics Committee of Hubei CDC (approval reference number: HBCDC-AF/SC-08/02.0). The demographical characteristics of the enrolled vaccinees were depicted in Table 1.

**Table 1.** Demographical characteristics of enrolled vaccinees

|  | **PLWH** | **HC** | **P value** |
| --- | --- | --- | --- |
| **Gender** |  |  |  |
| **Man (n)** | 29 | 16 | 0.3228 |
| **Woman (n)** | 13 | 12 |  |
| **Age (Year, mean±SD)** | 42.74±10.17 | 37.79±8.804 | 0.0392 |
| **BMI (Mean±SD)** | 23.97±6.967 | 24.07±3.8 | 0.2867 |

Note: PLWH, people living with HIV; HC, healthy control

### Peripheral lymphocyte count and plasma HIV-1 viral RNA detection

The peripheral lymphocyte counts and plasma HIV-1 viral RNA detections for HIV-1 infected individuals were performed using the BD Multitest™ CD3 FITC / CD8 PE / CD45 PerCP / CD4 APC reagent and the COBAS^®^ AmpliPrep / COBAS^®^ TaqMan^®^ HIV-1 Test (v2.0) kit, respectively. All the detections were conducted following the manufacturers’ instructions in the clinical laboratory of Shanghai Public Health Clinical Center.

### Titration of SARS-CoV-2 RBD binding antibody

In-house enzyme-linked immunosorbent assays (ELISA) were developed to measure SARS-CoV-2 RBD specific binding antibodies. High-binding 96-well EIA plates (Cat# 9018, Corning, USA) were coated with purified SARS-CoV-2 RBD protein (Cat# 40591-V08H, Sino Biological, China) at a final concentration of 1µg/ml in carbonate/bi-carbonate coating buffer (30mM NaHCO_3_,10mM Na_2_CO_3_, pH 9·6). Subsequently, the plates were blocked with 1×PBS containing 5% skimmed milk for 1 hour at 37°C. Next, 50μl of diluted human plasma was added to each well. After 1-hour incubation at 37°C, the plates were washed with 1×PBS containing 0·05% Tween20 for 5 times. Then, 50μl of an HRP labeled goat anti-human IgG antibody (Cat# ab6759, Abcam, UK) diluted in 1×PBS containing 5% skimmed milk were added to each well and incubated for 1 hour at 37°C. After a second round of wash, 50μl of TMB substrate reagent (Cat# MG882, MESGEN, China) was added to each well. 15 minutes later, the color development was stopped by adding 50μl of 1M H_2_SO_4_ to each well and the values of optical density at OD450_nm_ and OD630_nm_ were measured using 800 TS microplate reader (Cat# 800TS, Biotek, USA).

### Quantification of SARS-CoV-2 neutralizing antibodies

The neutralizing antibodies against SARS-CoV-2 were detected using a commercialized surrogate neutralization test developed by Suzhou Xinbo Biotechnology Ltd. Company (PerkinElmer, China). The rationale and method of this assay have been elaborated in previous studies.^11,12^ Briefly, plasma samples were firstly incubated with acridinium ester labeled SARS-CoV-2 RBD for 15 min at room temperature. Then, magnetic beads coated with purified human ACE2 protein were added into the mixture and incubated for 15 min. After washing, the acridinium ester labeled RBD bound with the magnetic beads were measured by a chemiluminescent reaction. The concentrations of SARS-CoV-2 neutralizing antibody correlate negatively with the luminescence intensities, which can be quantified by establishing a standard curve.

### Flowcytometry assays

To minimize measurement errors, freshly isolated PBMCs were preserved in liquid nitrogen until the last follow-up was completed. Upon recovery, the PBMCs were washed twice with R10 (RPMI1640 with 10% FBS). After counting, 1 million live (trypan blue negative) cells from each sample were added to 96-well round bottom plates and incubated with either R10 or R10 containing synthesized spike protein peptides (0.6μg/ml for each peptide) overnight. Then, the cells were washed and stained sequentially with live/dead dye(Zombie Aqua Fixable Viability Kit, cat#423101, BioLegend), surface marker antibodies (Anti-human-CD3-APC/cyanine 7, cat#300318, BioLegend; anti-human-CD4-BV605, cat#566148, BioLegend; anti-human-CD8-APC, cat#344722, BioLegend; anti-human-HLA-DR-BV421, cat#307636, BioLegend; anti-human-CD38-Percp-Cy5·5, cat#551400, BD Pharmingen) and an intracellular IFN-γ antibody (anti-human-IFN-γ-PE, cat#502509, BioLegend). Stained samples were analyzed using a BD Fortessa flow cytometer and data were analyzed with FlowJo software version 10 (TreeStar Inc., Ashland, OR, USA). The gating strategy is shown in Supplementary figure 1. The evaluation of T cell activation was based on the data generated from wells incubated with R10.

### Statistical analysis

All the statistical analyses were conducted using Graphpad Prism 9 (GraphPad Software, USA). Normality tests were performed before all downstream statistical analyses except the Chi square test. Comparisons between two groups were performed by the method of *t*-test. Differences among multiple groups were compared by the method of one-way ANOVA. The contingency analysis was done using the method of Chi-square test.

## Results

### Baseline characteristics of PLWH and HIV related events after vaccination

In this study, 28 healthy individuals and 42 PLWH were inoculated with two doses of the inactivated vaccine (BIBP-CorV) produced by Beijing Institute of Biological Products on 22 April 2021 and 25 May 2021, respectively. PLWH were slightly older than the control (42·74 ± 10·17 versus 37·79 ± 8·804 years, p=0·0392). The gender composition and the mean BMI were similar between PLWH and healthy individuals (Table 1). All the HIV-1 infected individuals have been on cART since being diagnosed with HIV-1 and have a peripheral CD4^+^ T cell count of >200 cells/μL. No solicited adverse reaction was observed among all participants. None of PLWH developed HIV-related clinical events during the study period from 22 April 2021 to 25 June 2021.

At the beginning of vaccination, 22 patients had an undetectable plasma viral load, 8 patients had detectable viral loads below 20 copies/ml and 12 patients had detectable viral loads beyond 20 copies/ml (Table 2). Unexpectedly, we found that the general viral burden among PLWH decreased after vaccination: 34 patients had an undetectable plasma viral load, 4 patients had detectable viral loads below 20 copies/ml and 4 patients had detectable viral loads beyond 20 copies/ml (P=0·0192) (Table 2). Meanwhile, our data also showed that the peripheral T cell counts decreased significantly after vaccination, but the CD4^+^/CD8^+^ T cell ratios remained stable (Table 2).

**Table 2.** Peripheral T cell counts and plasma viral loads of PLWH

|  | Baseline | 4 weeks post the 2<sup>nd</sup> dose | P value |
| --- | --- | --- | --- |
| **T cell count** |  |  |  |
| (Mean ± SD cells/μL, n) |  |  |  |
| CD3<sup>+</sup> T | 1550 ± 502, 28 | 1121 ± 336, 28 | Paired t-test, <0.0001 |
| CD4<sup>+</sup> T | 659 ± 222, 28 | 477 ± 151, 28 | Paired t-test, <0.0001 |
| CD8<sup>+</sup> T | 813 ± 361, 28 | 586 ± 240, 28 | Paired t-test, <0.0001 |
| **CD4<sup>+</sup> T/CD8<sup>+</sup> T ratio** |  |  |  |
| (Median, 25%-75% percentile, n) | 0.86, 0.60-1.05, 28 | 0.82, 0.62-1.25, 28 | Paired rank test, 0.5194 |
| **Plasma viral load** |  |  |  |
| >20 copies/mL (n) | 12 | 4 | Chi square test, 0.0192 |
| <20 copies/mL (n) | 8 | 4 |  |
| Undetectable (n) | 22 | 34 |  |

### The inactivated SARS-CoV-2 vaccine elicited comparable antibody and T cell responses in PLWH with those in healthy individuals

To evaluate the immunogenicity of the inactivated SARS-CoV-2 vaccine in our cohorts, peripheral blood samples were collected at baseline, 4 weeks after the first dose and 4 weeks after the second dose, respectively. The results of RBD binding antibody assays showed that the inactivated vaccine induced similar levels of RBD binding antibodies in the two cohorts after both the first and the second dose (Fig.1A). We found that the average fold increase of RBD binding antibody was significantly lower in PLWH (Fig.1B), which was presumably due to the relatively high background antibody levels in this cohort (Fig.1A). Neutralizing antibody responses were elicited in most participants after two doses of vaccination (Fig. 1D and 1E) and the magnitudes were similar between PLWH and the healthy control (Fig. 1F). In addition, we also found that the mean levels of S2 binding antibodies were similar between PLWH and healthy individuals both at baseline and 4 weeks post second vaccination (Supplementary Figure 2). The results of flowcytometry assays showed that two-dose inactivated SARS-CoV-2 vaccine induced significant T cell responses in both HC and PLWH (Fig.2). Similar magnitudes of S protein specific CD4^+^ (Fig. 2A) and CD8^+^ (Fig. 2B) T cell responses were observed in the two cohorts.

**Figure 1.**
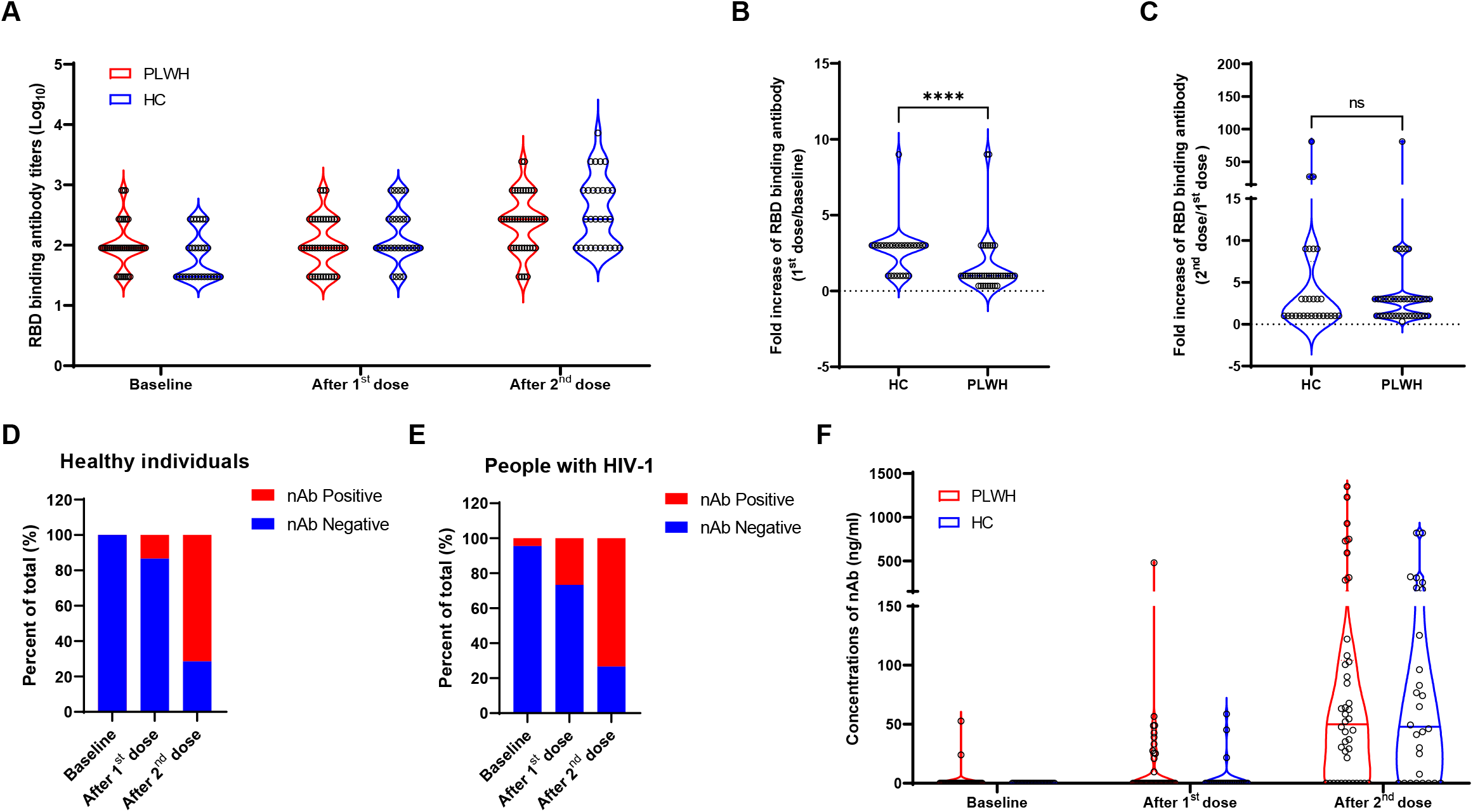
The inactivated SARS-CoV-2 vaccine elicited comparable RBD binding antibody and neutralizing antibody responses in PLWH with those in the control. All participants in our study received two doses of the inactivated SARS-CoV-2 vaccine (BIBP-CorV) produced by Beijing Institute of Biological Products at an interval of 4 weeks. Peripheral blood samples were collected at baseline, 4 weeks post the 1^st^ dose and 4 weeks post the 2^nd^ dose. RBD binding antibody (**A, B, C**) and neutralizing antibody (**D, E, F**) responses were measured and compared between PLWH (n=28) and healthy individuals (n=42). Statistical analyses were performed by the method of non-parametric t test. ****, P<0·0001.

**Figure 2.**
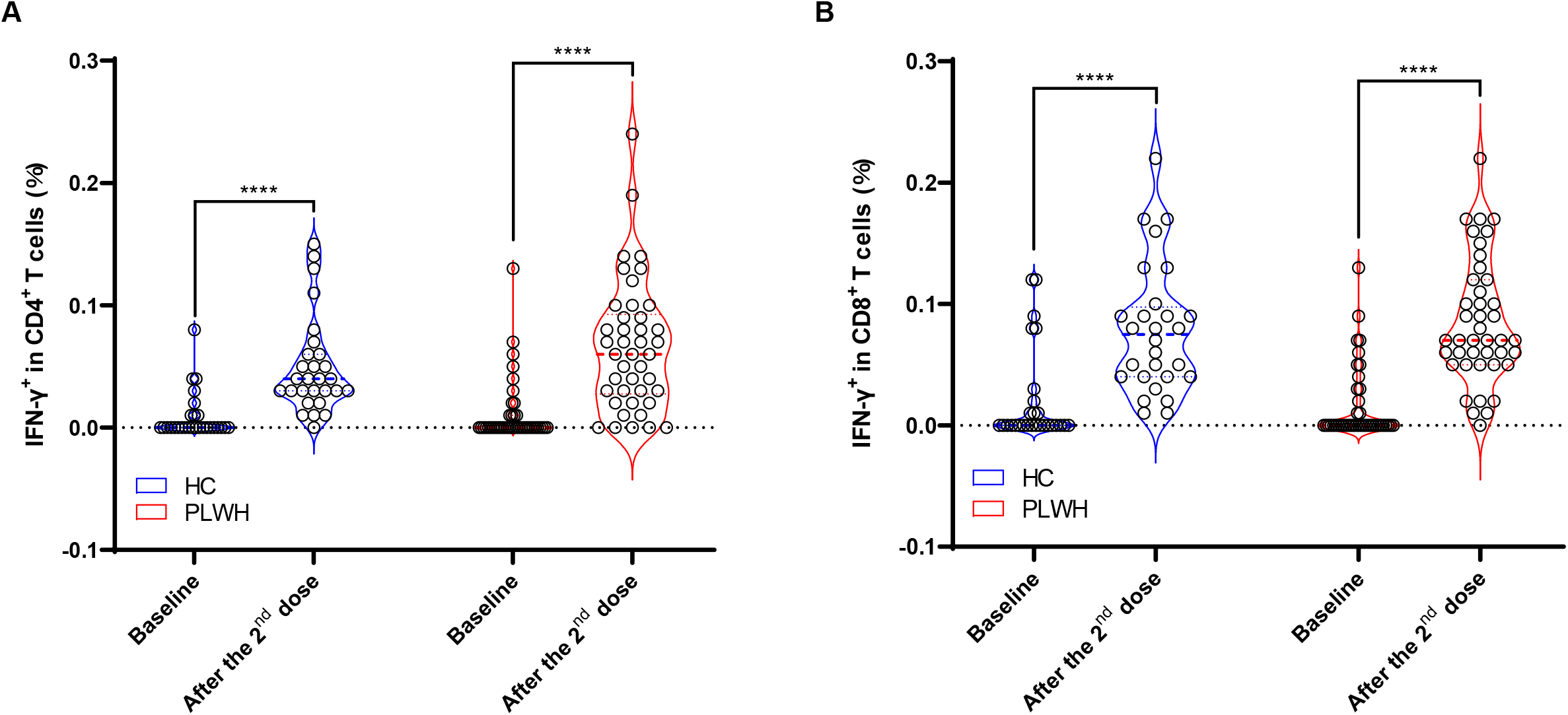
Comparisons of S protein specific T cell responses between PLWH and healthy individuals. S protein specific IFN-γ secreting T cells were measured using flowcytometry. The inactivated vaccine elicited similar levels of S protein specific CD4^+^ (**A**) and CD8^+^ T cell responses (**B**) in PLWH and HC. Statistical analyses were performed by the method of non-parametric t test. ****, P<0·0001.

### Baseline CD4/CD8 ratios were associated with antibody responses after the vaccination in the cohort of PLWH

To understand whether the baseline immune status can influence the vaccine induced antibody responses in PLWH, we further analyzed the relationships between baseline T cell counts and post vaccination antibody responses. Our data showed that neither the CD4^+^ T cell counts nor the CD8^+^ T cell counts at baseline correlated with the vaccine induced antibody responses (Data not shown). While, the CD4^+^/CD8^+^ T cell ratios at baseline were found to be associated with antibody responses after the second dose of vaccination (Fig.3). More specifically, the average RBD binding and neutralizing antibody responses of HIV-1 positive vaccinees with low baseline CD4^+^/CD8^+^ T cell ratios (<0·6) were significantly lower than those with medium (0·6-1·0) or high (≥1·0) baseline CD4^+^/CD8^+^ T cell ratios (Fig.3A and 3B). Slightly different from the RBD binding antibody and the nAb responses, we observed that S2 binding antibody responses in HIV-1 positive vaccinees with medium baseline CD4^+^/CD8^+^ T cell ratios were significantly higher than those with both low and high baseline CD4^+^/CD8^+^ T cell ratios (Fig.3C). We also analyzed the impacts of baseline CD4^+^/CD8^+^ T cell ratios on T cell responses and found that there was no significant difference among PLWH with low, medium and high baseline CD4^+^/CD8^+^ T cell ratios (Supplementary figure 3).

**Figure 3.**
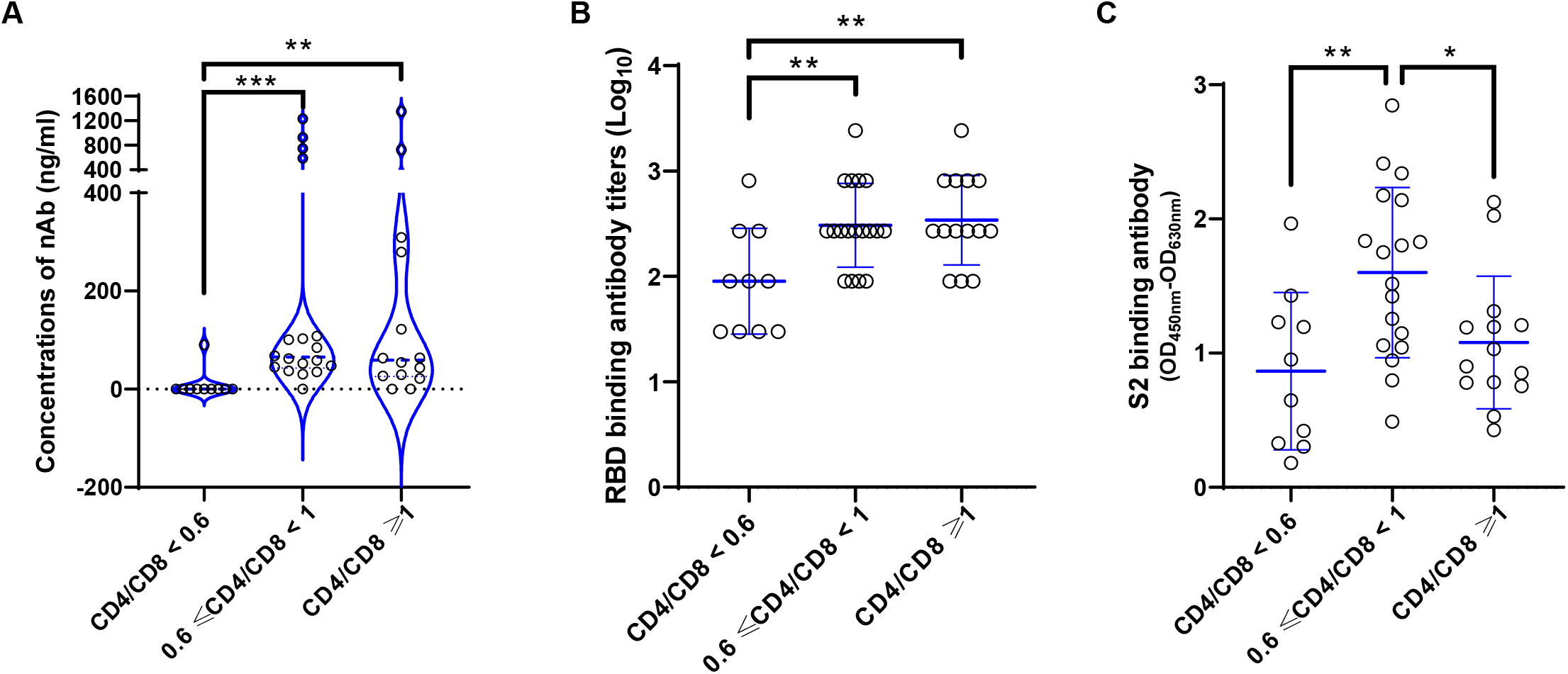
Baseline CD4^+^/CD8^+^ T cell ratios were associated with specific antibody responses after vaccination in PLWH. HIV-1 infected participants were stratified into 3 subgroups according to their baseline CD4^+^/CD8^+^ T cell ratios (<0·6, 0·6∼1·0 and ≥1·0). RBD binding antibody (**A**), neutralizing antibody (**B**) and S2 binding antibody (**C**) responses were compared among the three subgroups. Statistical analysis for (**A**) was performed by the method of non-parametric t test and statistical analyses for (**B**) and (**C**) were done by the method of parametric t test. *, P<0·05; **, P<0·01; ***, P<0·001.

### The patterns of vaccine induced T cell activation were different between HC and PLWH

Vaccine induced activation of T cells was considered as a major constraint that might compromise vaccine conferred benefits in PLWH.^13-15^ To investigate whether the vaccine induced T cell activation was associated with the observed fluctuation of HIV-1 viral load, we measured the expressions of CD38 and HLA-DR on CD4^+^ and CD8^+^ T cells by flowcytometry. We observed that the patterns of vaccine induced T cell activation were different between HC and PLWH (Fig.4). Of note, our data showed that the percentages of CD38^+^HLA-DR^-^ CD4^+^ T cells (Fig.4B) increased, while the percentages of CD38^-^HLA-DR^+^ CD8^+^ T cells (Fig.4D) decreased in PLWH after vaccination. The decrease of CD38^-^HLA-DR^+^CD8^+^ T cells has been reported by a previous study,^13^ however, the underlying mechanism is not clear. We further analyzed the association between the observed T cell activation and plasma viral load fluctuation. Although no statistical significance was reached, our results showed that the vaccine induced fold-changes of CD38^+^HLA-DR^-^, CD38^-^HLA-DR^+^ and CD38^+^HLA-DR^+^CD4^+^ T cells tended to be higher in viral increased PLWH (Supplementary figure 4).

**Figure 4.**
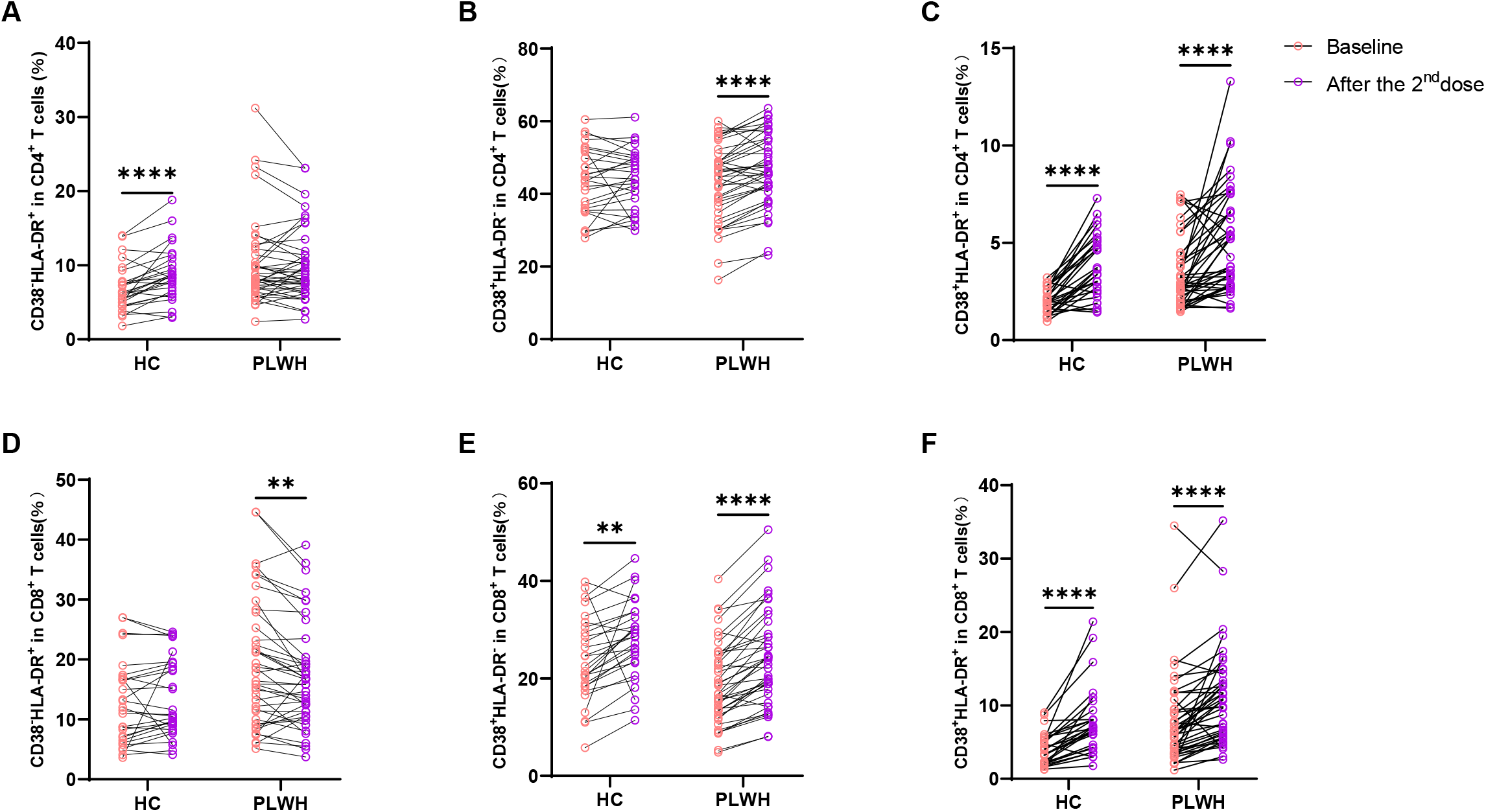
The patterns of vaccine induced T cell activation were different between PLWH and HC. The activation of T cells was assessed through detecting the expressions of CD38 and HLA-DR on T cells. (**A, B, C**) The percentages of CD38^-^HLA-DR^+^, CD38^+^HLA-DR^-^ and CD38^+^HLA-DR^+^ among CD4^+^ T cells. (**D, E, F**) The percentages of CD38^-^HLA-DR^+^, CD38^+^HLA-DR^-^ and CD38^+^HLA-DR^+^ among CD8^+^ T cells. Statistical analyses were performed by the method of two-way ANOVA. **, P<0·01; ****, P<0·0001.

## Discussion

Even if the replication of HIV is controlled by cART, it can still be harder for PLWH to defend against infectious diseases.^16^ Therefore, vaccination is recommended for PLWH whose peripheral CD4^+^ T cell counts are more than 200 cells/μL.^17^ However, due to the compromised immune status, the vaccine elicited immune responses might be impaired in PLWH.^18-21^ The efficacies of COVID-19 vaccines in PLWH were also seriously concerned, because it has been suggested that HIV-1 infection could alter the host immune responses against SARS-CoV-2.^22^ Here, we provided evidence showing that an inactivated SARS-CoV-2 vaccine (BIBP-CorV) was immunogenic and safe in PLWH. Importantly, our data demonstrated that the inactivated vaccine induced comparable antibody and T cell responses between PLWH and HC.

All the HIV-1 positive participants in our cohort had a CD4^+^ T cell count of >200 cells/μL both at baseline and 4 weeks post vaccination. We found a statistically significant decrease in CD4^+^ T cell count between the baseline levels and those measured following the second vaccination. The decrease of CD4^+^ T cell count after vaccination had also been observed in PLWH inoculated with the BNT162b2 mRNA vaccine and several other vaccines usually recommended for PLWH.^23,24^ Increases in activated T cells were found to be associated with decreases in CD4^+^ T cell count among PLWH received vaccination,^24^ but the reason is still not clear. Of note, both our and other studies did not identify any adverse clinical manifestation that was associated with the drop of CD4^+^ T cell count.^23,24^ Stable cART might be crucial to counteract the potential vaccine related deleterious effect in PLWH according to a recently published case report, which observed viral activation and CD4^+^ T cell loss in a treatment-naïve HIV-positive patient after receiving two doses of inactivated COVID-19 vaccines.^25^ Quite surprisingly, we observed a significant drop of general viral burden in the PLWH cohort after vaccination. As percentages of activated T cells may correlate with HIV-1 replication, we further evaluated the vaccine induced T cell activation by measuring the expressions of CD38 and HLA-DR on T cells. We found that the percentages of CD38^+^HLA-DR^-^CD4^+^ T cells and CD38^+^HLA-DR^+^CD4^+^ T cells increased significantly after vaccination. We speculate that the elimination of activated HIV-1 infected CD4^+^ T cells by cART or autologous CTLs might account for the observed drop of HIV burden in the PLWH cohort.

Despite the generally comparable antibody responses between PLWH and healthy individuals, we demonstrated that HIV-1 infected vaccinees with low baseline CD4^+^/CD8^+^ T cell ratio (<0·6) generated weaker antibody responses after vaccination, which was in line with previous reports showing that CD4^+^/CD8^+^ T cell ratios could predict vaccine induced antibody responses in HIV-1 infected patients and HIV negative elderly.^26,27^ Additionally, we showed that the baseline CD4^+^/CD8^+^ T cell ratios did not impact the induction of specific T cell responses. Given that most vaccine induced antibody responses are CD4^+^ T cell dependent, a potential explanation for the above phenomena is that the low CD4^+^/CD8^+^ T cell ratio might reflect the suboptimal support for antibody responses.

A major limitation of this study is the inadequate follow-up, which do not allow us to compare the persistence of antibody responses between PLWH and the healthy control. Despite of this limitation, our study demonstrates that the inactivated COVID-19 vaccine was immunogenic and safe in PLWH stable on cART.

## Data Availability

The data that support the findings of this study are available from the corresponding author upon reasonable request.

## Author contributions

YW and HT were involved in the study design and supervision; YF, YZ, ZH, XT GW, DC and LJ were involved in data collection; YR, WW, JW, LS, WZ and YW performed the data analysis; YW, YF and HT drafted the manuscript; WZ, LS, YW and HT revised the manuscript. HH coordinated the enrollment of PLWH. The underlying data have been verified by YF and YW.

## Data sharing

Anonymized participant-level data will be made available 3 months following publication, upon requests directed to the corresponding author. After being approved by the sponsors (Fudan University and Hubei provincial CDC) and investigators, the data can be shared through a secure online platform. All data will be made available for a minimum of 5 years after publication.

## Acknowledgments

We thank Dr. Lianguo Ruan from Wuhan Jinyintan Hospital for his help in PLWH cohort enrollment. We thank the volunteers who participated in this study. This work was funded by the National Natural Science Foundation of China (Grant No. 81971559, 82041010).

