## Supplements for "Immunogenicity and safety of an inactivated SARS-CoV-2 vaccine in people living with HIV-1"

### Supplementary materials


| Contents | Page numbers |
|---|---|
| Supplementary figure legends | 1 |
| Supplementary figure 1: The gating strategy of flow cytometry | 2 |
| Supplementary figure 2: Comparisons of S2 binding antibody responses between PLWH and healthy individuals | 3 |
| Supplementary figure 3: Baseline CD4/CD8 ratios were not significantly associated with the vaccine elicited T cells responses in PLWH | 4 |
| Supplementary figure 4: The vaccine induced activation of CD4+ T cells in individuals with elevated viral loads tended to be higher than those in viral load decreased individuals | 5 |

#### Supplementary figure legends

**Supplementary Figure 1 The gating strategy of flow cytometry**

**Supplementary Figure 2 Comparisons of S2 binding antibody responses between PLWH and healthy individuals**

The SARS-CoV-2 S2 specific binding antibodies were detected using an in-house ELISA method. The inactivated COVID-19 vaccine elicited similar levels of S2 binding antibodies in PLWH and HC. Statistical analyses were performed by the method of non-parametric t test. ****, P<0·0001.

**Supplementary Figure 3 Baseline CD4/CD8 ratios were not significantly associated with the vaccine elicited T cells responses in PLWH**

HIV-1 infected participants were stratified into 3 subgroups according to their baseline $CD4^+/CD8^+$ T cell ratios (<0·6, 0·6~1·0 and ≥1·0). S protein specific IFN-γ secreting $CD4^+$ (**A**) and $CD8^+$ T cells (**B**) were compared among the three subgroups. Statistical analyses were performed by the method one-way ANOVA.

**Supplementary Figure 4 The vaccine induced activation of $CD4^+$ T cells in individuals with elevated viral loads tended to be higher than those in viral load decreased individuals**

Among the PLWH cohort, 14 participants were found with decreased HIV-1 viral loads and 5 participants showed increased HIV-1 viral loads after two doses of vaccination. The activation levels of $CD4^+$ T cells were compared between these two subgroups. (**A**) The fold changes (After the $2^{nd}$ vaccination/baseline) of $CD38^-HLA\text{-}DR^+CD4^+$ T cell percentages. (**B**) The fold changes of $CD38^+HLA\text{-}DR^-CD4^+$ T cell percentages. (**C**) The fold changes of $CD38^+HLA\text{-}DR^+CD4^+$ T cell percentages.

**Supplementary Figure 1 The gating strategy of flow cytometry**

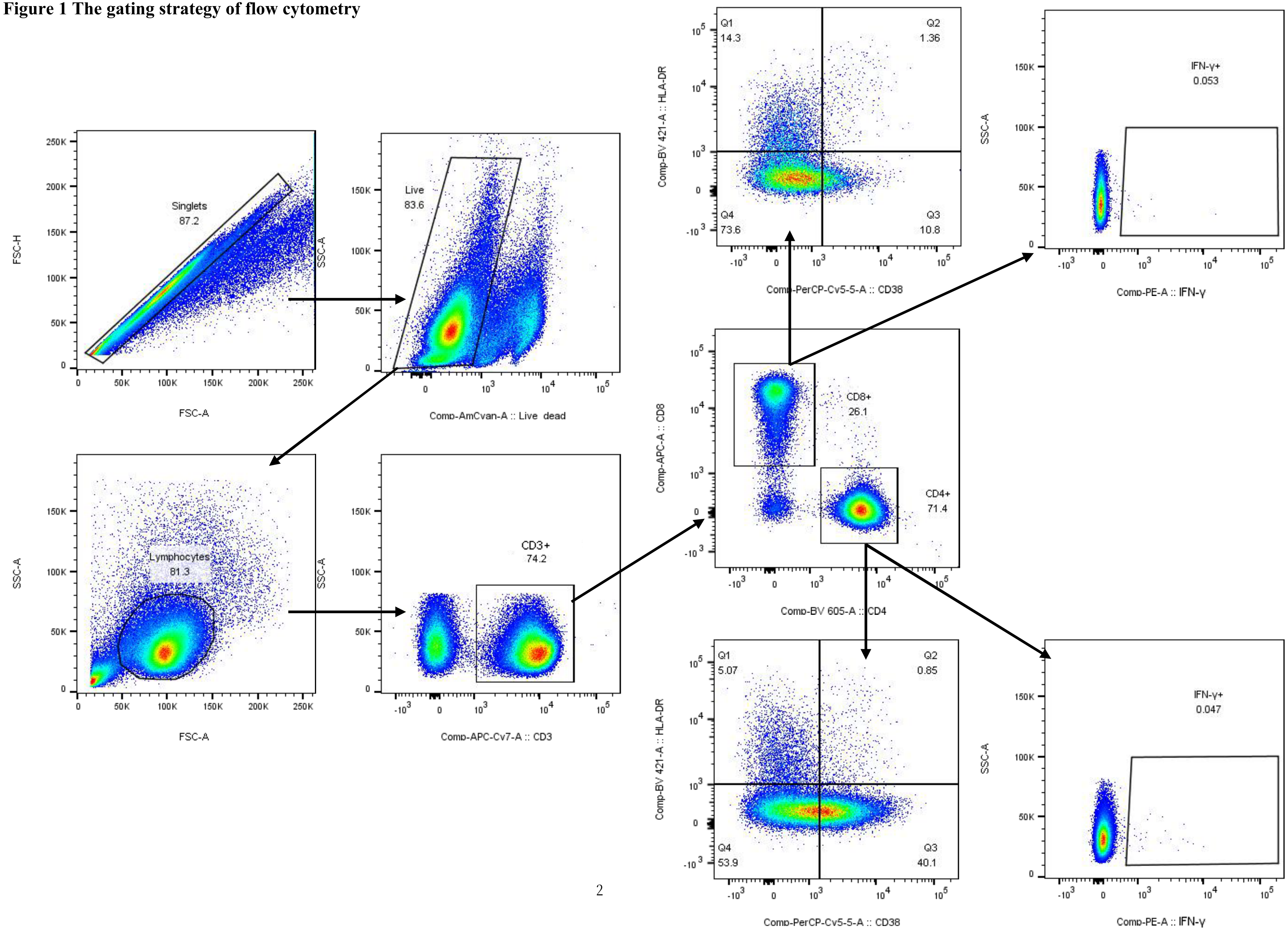

**Supplementary Figure 2 Comparisons of S2 binding antibody responses between PLWH and healthy individuals**

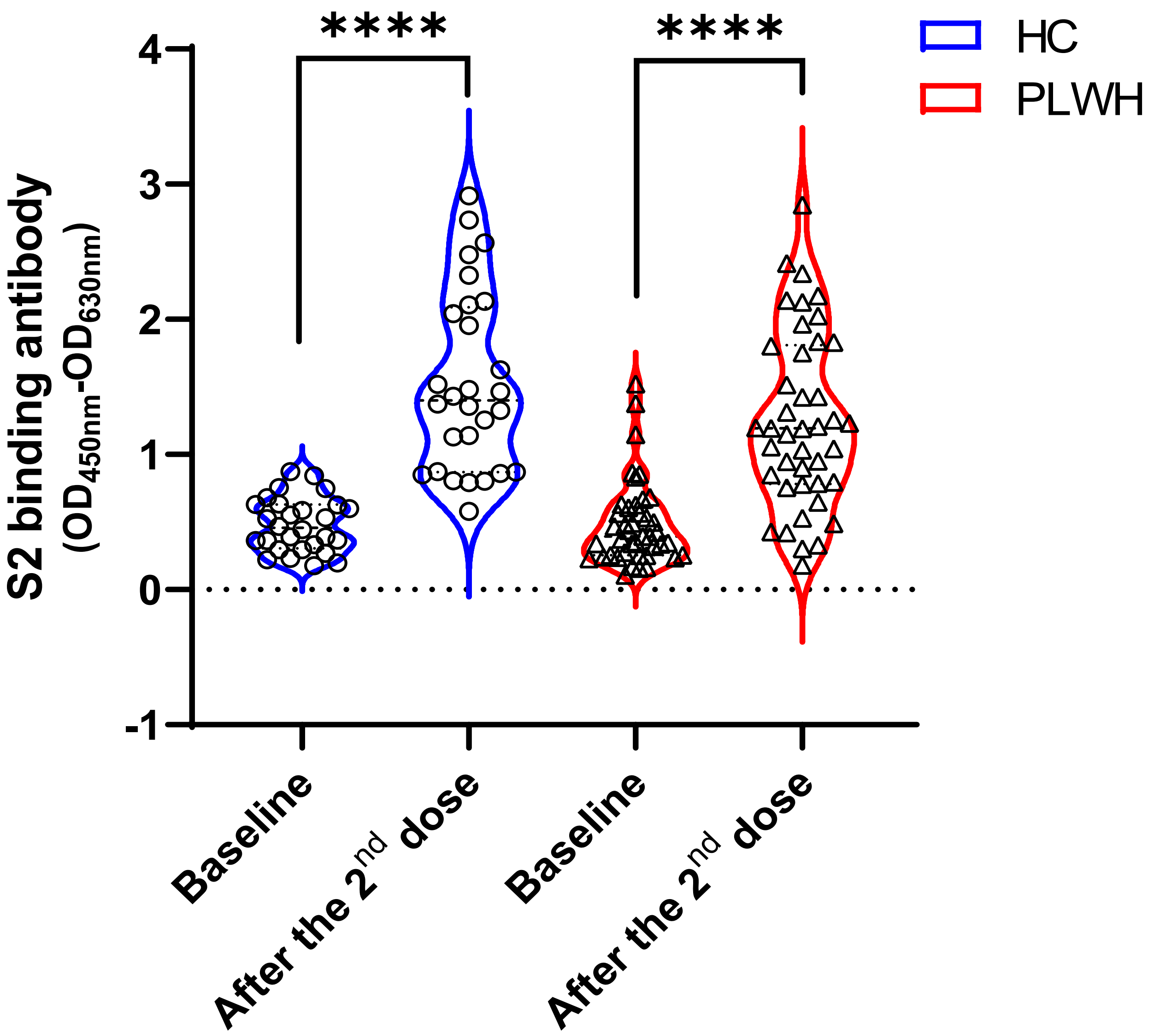

**Supplementary Figure 3 Baseline CD4/CD8 ratios were not significantly associated with the vaccine elicited T cells responses in PLWH**

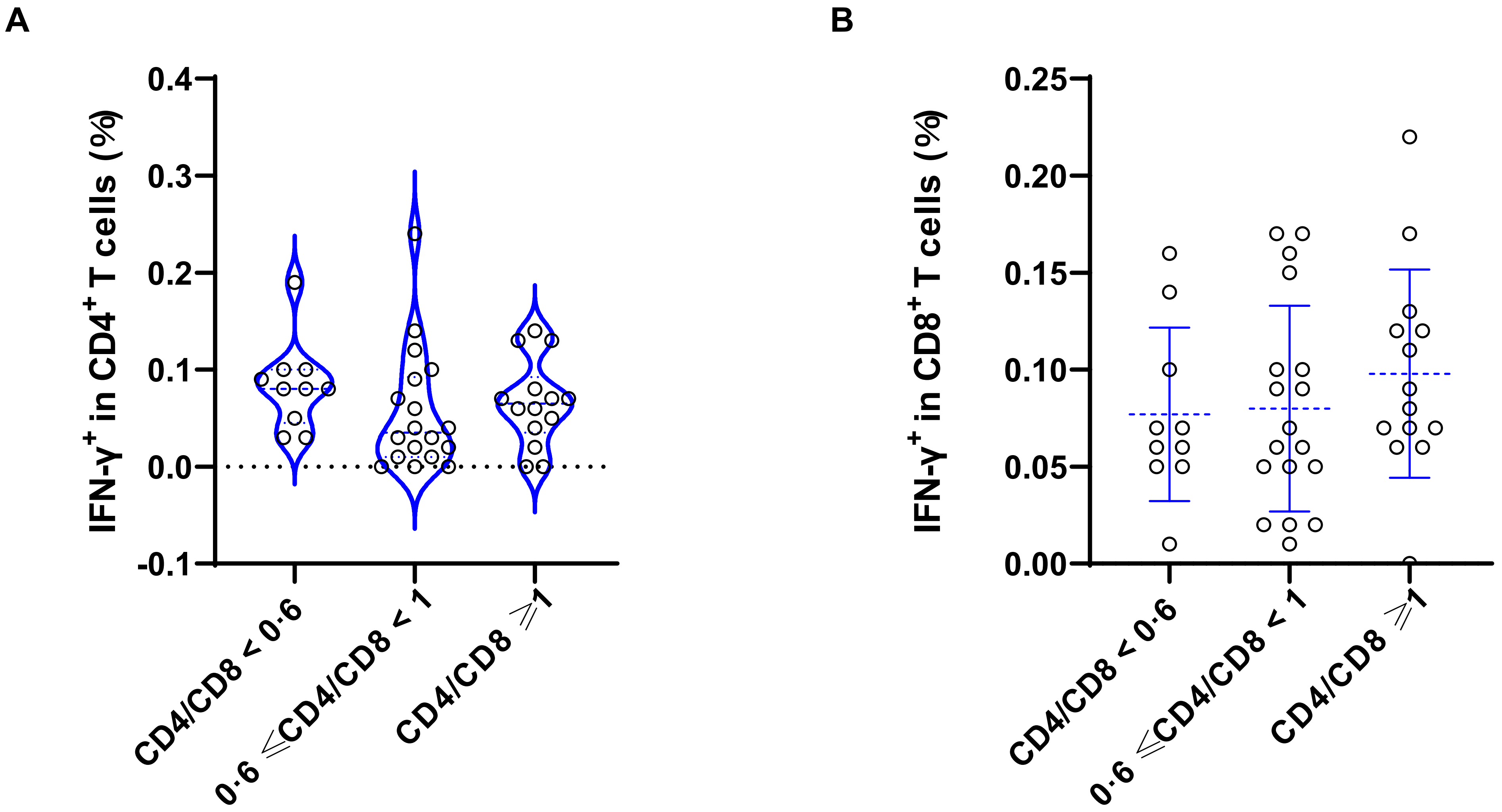

**Supplementary Figure 4 The vaccine induced activation of CD4$^{+}$ T cells in individuals with elevated viral loads tended to be higher than those in viral loads decreased individuals**

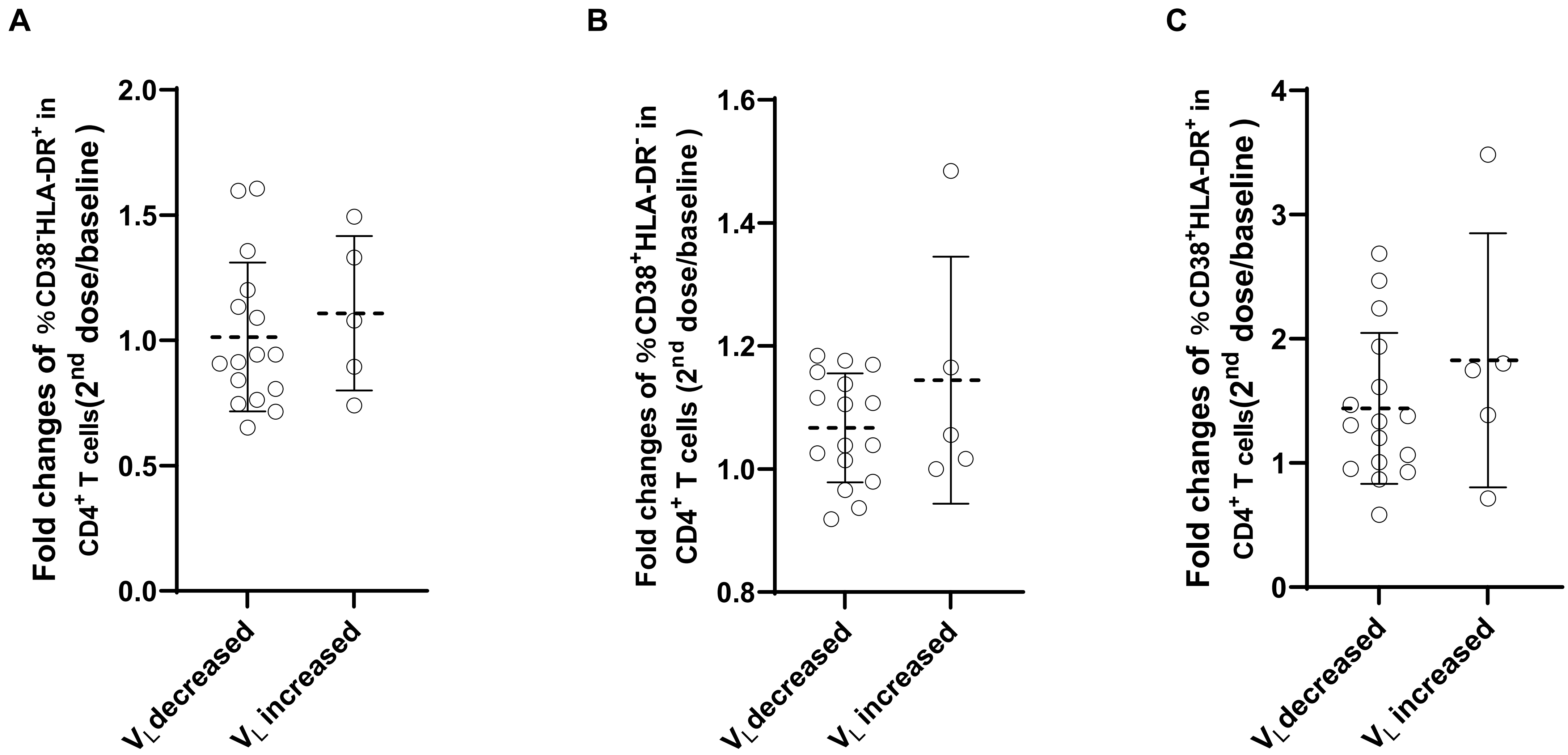
